# Reporting of socio-demographic characteristics of trial participants in infectious diseases clinical trials – a systematic review

**DOI:** 10.1101/2025.03.16.25324074

**Authors:** Sean W. X. Ong, Christina Blagojevic, Aliya Bryce, Aaron Ovadia, Matthew Slater, Daire Pryal, Rodrigo Escobar Careaga, Hadrien Moffroid, Arvind Yerramilli, Esmita Charani, Nick Daneman, Steven Y. C. Tong

**Affiliations:** Institute of Health Policy, Management and Evaluation, University of Toronto, Toronto, Canada; Department of Infectious Diseases, University of Melbourne, at the Peter Doherty Institute for Infection and Immunity, Melbourne, Australia; Sunnybrook Health Sciences Centre, Toronto, Canada; Victorian Infectious Diseases Service, Royal Melbourne Hospital, Peter Doherty Institute for Infection and Immunity, Melbourne, Australia; University of Toronto, Toronto, Canada; Faculty of Medicine, Dentistry and Health Sciences, University of Melbourne, Melbourne, Australia; Division of Infectious Diseases & HIV Medicine, Department of Medicine, Groote Schuur Hospital, University of Cape Town, Cape Town, South Africa; Faculty of Health and Medical Sciences, University of Liverpool, Liverpool, United Kingdom

**Author notes:** **Corresponding Author:** Dr Sean W. X. Ong, Institute of Health Policy, Management and Evaluation, University of Toronto, Toronto, Canada. These two senior authors contributed equally (Nick Daneman, Steven Y. C. Tong).

**Keywords:** clinical trials, representativeness, social determinants of health, demographics, reporting quality, infectious diseases

## Abstract

**Background:** Reporting of demographic characteristics in randomised clinical trials (RCT) is recommended to facilitate assessment of generalisability to other populations. However, there is a lack of consensus as to what variables should be reported, and there are limited data describing current research practice.

**Methods:** We conducted a systematic review of all infectious diseases-related RCTs published between January 2014 to August 2023 in ten selected high impact journals. Outcomes of interest were the reporting of five patient-level sociodemographic characteristics, as recommended by the CONSORT-Equity 2017 extension to the CONSORT reporting guidelines: (a) ethnicity, (b) sex and/or gender, (c) education level, (d) socioeconomic status (SES), and (e) rurality. We summarised descriptive results for the reporting of each characteristic overall, by trial type (health equity-related vs non-health equity-related), subject area, and year of publication. We fitted multivariable logistic regression models to identify trial characteristics associated with the reporting of each characteristic.

**Results:** 4234 articles were screened and 1343 were included. Almost all trials (97.4%) reported sex and/or gender. In contrast, less than half (49.3%) reported ethnicity, and only a minority reported education level (9.0%), SES (9.0%), and rurality (3.9%). There was no improvement in reporting of each characteristic over the 10-year period. Subject area, funding source, whether a trial was health-equity related, use of a medical writer, and trial setting (high vs low/middle-income country) were significantly associated with the reporting of ethnicity, education level, and SES.

**Conclusion:** Reporting of socio-demographic characteristics in infectious diseases RCTs is inconsistent and has not improved over time.

## BACKGROUND

Reporting of baseline characteristics of participants in a randomized clinical trial (RCT), often summarized in the form of a “Table 1”, allows readers to judge the relevance of trial results to other patient populations, i.e., whether the trial results have generalizability or external validity [1]. These often include demographic characteristics as well as other clinically-relevant information such as comorbidities or disease factors. Often only age and sex are reported [2], even though it is well established that numerous socio-demographic characteristics impact health behaviours and outcomes, including access to participation in clinical trials [3-6]. This is especially relevant in infectious diseases, where socio-demographic characteristics are associated with both the likelihood of acquiring infection and the severity of ensuing infection [7-11].

**Table 1:** Summary of characteristics of included trials.

| Characteristic | Number (%) |
| --- | --- |
| Journal |  |
| *Annals of Internal Medicine* | 35 (2.6) |
| *British Medical Journal* | 31 (2.3) |
| *Clinical Infectious Diseases* | 298 (22.2) |
| *Clinical Microbiology & Infection* | 58 (4.3) |
| *Journal of the American Medical Association* | 117 (8.7) |
| *Journal of Infection* | 19 (1.4) |
| *Journal of Infectious Diseases* | 123 (9.2) |
| *The Lancet* | 171 (12.7) |
| *Lancet Infectious Diseases* | 241 (17.9) |
| *New England Journal of Medicine* | 250 (18.6) |
| Year |  |
| 2014 | 132 (9.8) |
| 2015 | 134 (10.0) |
| 2016 | 120 (8.9) |
| 2017 | 86 (6.4) |
| 2018 | 105 (7.8) |
| 2019 | 130 (9.7) |
| 2020 | 130 (9.7) |
| 2021 | 218 (16.2) |
| 2022 | 166 (12.4) |
| 2023 | 122 (9.1) |
| Funding source |  |
| Industry-funded | 331 (24.7) |
| Investigator-initiated | 223 (16.6) |
| Mixed funding sources | 772 (57.6) |
| Not available / not reported | 15 (1.1) |
| Subject area |  |
| Antimicrobial stewardship and infection control | 51 (3.8) |
| Bacterial and fungal infections | 237 (17.6) |
| COVID-19 | 147 (10.9) |
| Critical care | 37 (2.8) |
| HIV and STIs | 190 (14.1) |
| Tuberculosis | 61 (4.5) |
| Tropical medicine | 174 (13.0) |
| Vaccines | 311 (23.2) |
| Viral infections (excluding COVID-19) | 135 (10.1) |
| Sample size, median (IQR) | 465 (182–1274) |
| Number of centres, median (IQR) | 8 (2–31) |
| Trial setting |  |
| High-income country | 668 (50.1) |
| Low or middle-income country | 474 (35.5) |
| Mixed (high and low/middle-income country) | 192 (14.4) |
| Health equity-related | 216 (16.1) |
| Use of a medical writer | 297 (22.1) |

COVID-19 = coronavirus disease 2019; HIV = human immunodeficiency virus; STI = sexually transmitted infection; IQR = interquartile range

The CONSORT-Equity 2017 extension to the CONSORT reporting guidelines suggests that for health equity-related RCTs (defined as RCTs that evaluate an intervention focused on people experiencing social disadvantage or that explore different effects of an intervention between groups with varying levels of social disadvantage), baseline characteristics should be reported across relevant socio-demographic characteristics [12]. The original CONSORT 2010 guidelines for clinical trials did not provide guidance on what baseline characteristics should be reported, potentially contributing to inconsistent reporting across trials [13].

Whilst gaps in reporting of socio-demographic variables in published RCTs have been described [14-16], there are limited data reviewing the extent of reporting of these socio-demographic characteristics specifically in infectious diseases RCTs. We aimed to conduct a systematic review to evaluate the reporting of socio-demographic characteristics of trial participants in infectious diseases RCTs and identify gaps in current research practice.

## METHODS

### Objective

The primary objective was to describe reporting of five socio-demographic characteristics in infectious diseases clinical trials published over a ten-year period (2014 to 2023). The socio-demographic characteristics of interest were selected from the PROGRESS-Plus framework, and consisted of: (1) ethnicity, (2) sex and/or gender, (3) education level, (4) socioeconomic status, (5) rurality or geographical location.[17] The PROGRESS-Plus framework outlines characteristics that influence health opportunities and outcomes, and is the framework recommended by the CONSORT-Equity 2017 extension to guide identification of which socio-demographic variables should be reported [12].

### Search strategy and eligibility criteria

This systematic review was conducted according to a pre-specified protocol registered on PROSPERO (CRD42023450864) and conducted in accordance with PRISMA guidelines [18]. The Ovid MEDLINE database was searched for all English-language manuscripts published between 01 January 2014 to 01 August 2023 in ten selected high impact journals that publish RCTs (five general medical journals and five infectious diseases specific journals). The full search strategy is shown in the **Supplementary Appendix**. Inclusion criteria were: (a) papers reporting primary results of an RCT, as defined as a prospective study evaluating an intervention with at least two intervention groups, with randomization occurring either at the individual patient or cluster level, and (b) the primary disease of interest is an infectious disease or syndrome, defined as a disease or syndrome caused by bacteria, viruses, fungi, parasites, or prions. Exclusion criteria were: (a) no full manuscript published, (b) study protocol only without trial results, (c) meta-analyses combining multiple trial results, (d) papers reporting secondary results or analyses of clinical trials that already have had primary results reported (e.g., longer-term outcomes, secondary sub-studies of a primary RCT), and (e) no individual-level baseline characteristics reported (e.g., in a cluster-randomized trial only reporting cluster-level characteristics).

### Study screening and data collection

Search results were imported into Covidence and de-duplicated. Title and abstract screening was conducted by two independent investigators (SWXO, CB) to assess suitability for inclusion, and any disagreements were resolved through discussion. Data extraction was conducted using a standardized data collection form by a team of nine data extractors. To ensure quality of data extraction, each data extractor had to first complete standardized training and a pilot extraction of 20 studies which was cross-referenced to a duplicate standardized extraction. Concordance of collected fields had to be >95% before extractors were permitted to complete data extraction independently; failing resulted in re-training and a repeat extraction of an additional 20 studies that was cross-referenced again before permitting independent extraction.

The outcomes of interest were the reporting of five selected socio-demographic variables of interest: (1) ethnicity, (2) sex and/or gender, (3) education level, (4) socioeconomic status, (5) rurality or geographical location. Detailed definitions of each variable are provided in the **Supplementary Methods**. Each outcome of interest was treated as a binary outcome (either presence or absence of reporting). Reporting had to be quantitative to be considered as presence of reporting (e.g., number of participants with or without proportion), and could be either in the summary table, main results text, or supplementary information. If a socio-demographic variable was part of the trial design, population definition, or eligibility criteria (e.g., a human papilloma virus vaccine trial conducted amongst women only, or a malaria chemoprevention trial conducted in rural villages only), the reporting of that specific variable would be considered “not applicable” (e.g., sex/gender and rurality respectively for the two stated examples, since these are implicit in the study design and hence does not need to be explicitly reported), and these studies would be excluded from the analysis for that particular variable.

In addition to reporting of socio-demographic variables, we recorded multiple trial characteristics of interest: year of publication, funding type (industry-sponsored, investigator-initiated, or mixed), phase of trial, trial design, subject area, sample size, number of centres, country where study was conducted, primary setting, health equity relatedness, and use of a professional medical writer.

### Statistical analysis

We calculated proportions of trials reporting each individual socio-demographic variable of interest, using a denominator of all trials, excluding those that were flagged “not applicable” for that variable (because the trial population was defined based that variable, as explained above). Proportions were calculated for the entire set of trials, and separately stratified by health equity relatedness, year of publication, and subject area. We also fitted a multivariable logistic regression model to identify study characteristics independently associated with reporting for each socio-demographic variable, with reporting of the variable of interest as the dependent variable, and the following trial characteristics as independent variables: year of publication (2013 – 2017 vs 2018 – 2023), subject area, sample size, number of centres, funding source, health equity relatedness, use of a professional medical writer, and income level of the country/countries where the trial was conducted. 2017 was taken as the cut-off for dichotomizing year of publication; the CONSORT-Equity statement was published that year. Countries were divided into high-income vs low/middle-income based on classification from the World Bank as of 2024.[19] Visualizations and data analysis were performed using R (version 4.4.1, R Foundation for Statistical Computing, Vienna, Austria).

As the objective of this study was to assess reporting of socio-demographic variables of interest, rather than appraise the results of RCTs, we did not conduct risk of bias assessment, certainty assessment, or assessment of study heterogeneity.

## RESULTS

4234 studies were screened, of which 1454 were evaluated by full text for eligibility. A further 111 were excluded, resulting in 1343 studies included in the final analysis (PRISMA study flow diagram in **Figure 1**). Study characteristics are summarized in **Table 1**. There was a good distribution of trials across the 10 included journals, 10 years of publication within the study period, and 9 subject area categories. 216 (16.1%) trials were identified as health equity-related (i.e., they should be reporting relevant socio-demographic characteristics according to CONSORT-Equity recommendations).

**Figure 1:**
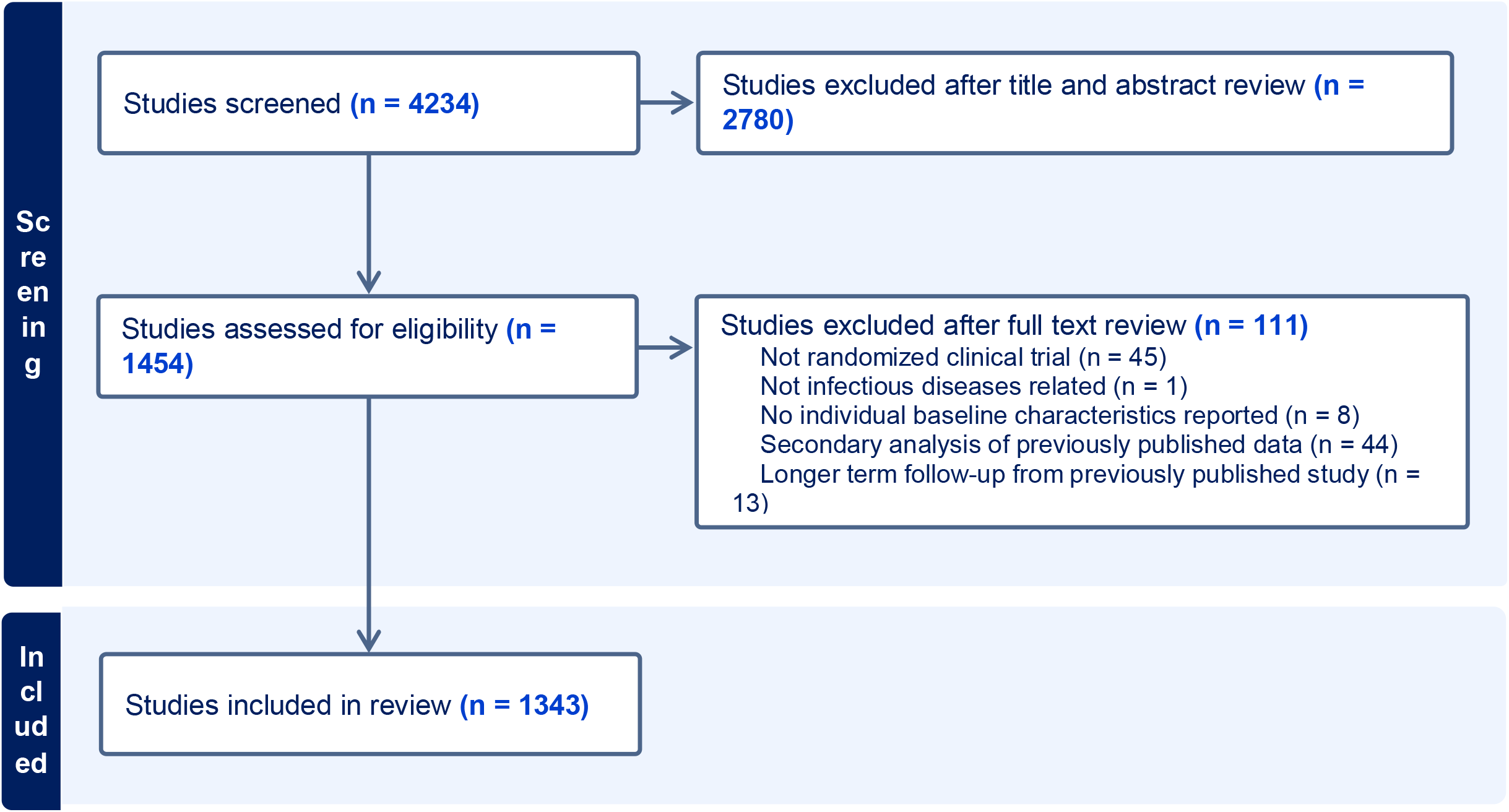
PRISMA study flow diagram.

Almost all trials (1201 of 1233; 97.4%) reported sex and/or gender. In contrast, less than half (654 of 1326; 49.3%) reported ethnicity, and only a minority reported education level (113 of 1252; 9.0%), socioeconomic status (120 of 1340; 9.0%), and rurality (45 of 1269; 3.9%) (**Figure 2A**). 63 (4.7%) trials did not report any of the socio-demographic variables of interest, 559 trials (41.6%) reported one variable, 621 trials (46.2%) reported two, 68 trials (5.1%) reported three, 31 trials (2.3%) reported four, while only 1 (0.07%) trial reported all five variables. There was also variation in how each socio-demographic variable was reported (**Supplementary Table 1**). For example, of the 654 trials that reported ethnicity, 117 (17.9%) reported only binary categories (e.g. white and non-white), while 537 (82.1%) reported multiple possible categories. There was wide variation in how education level, socioeconomic status, and rurality variables were reported (**Supplementary Table 1**).

**Figure 2:**
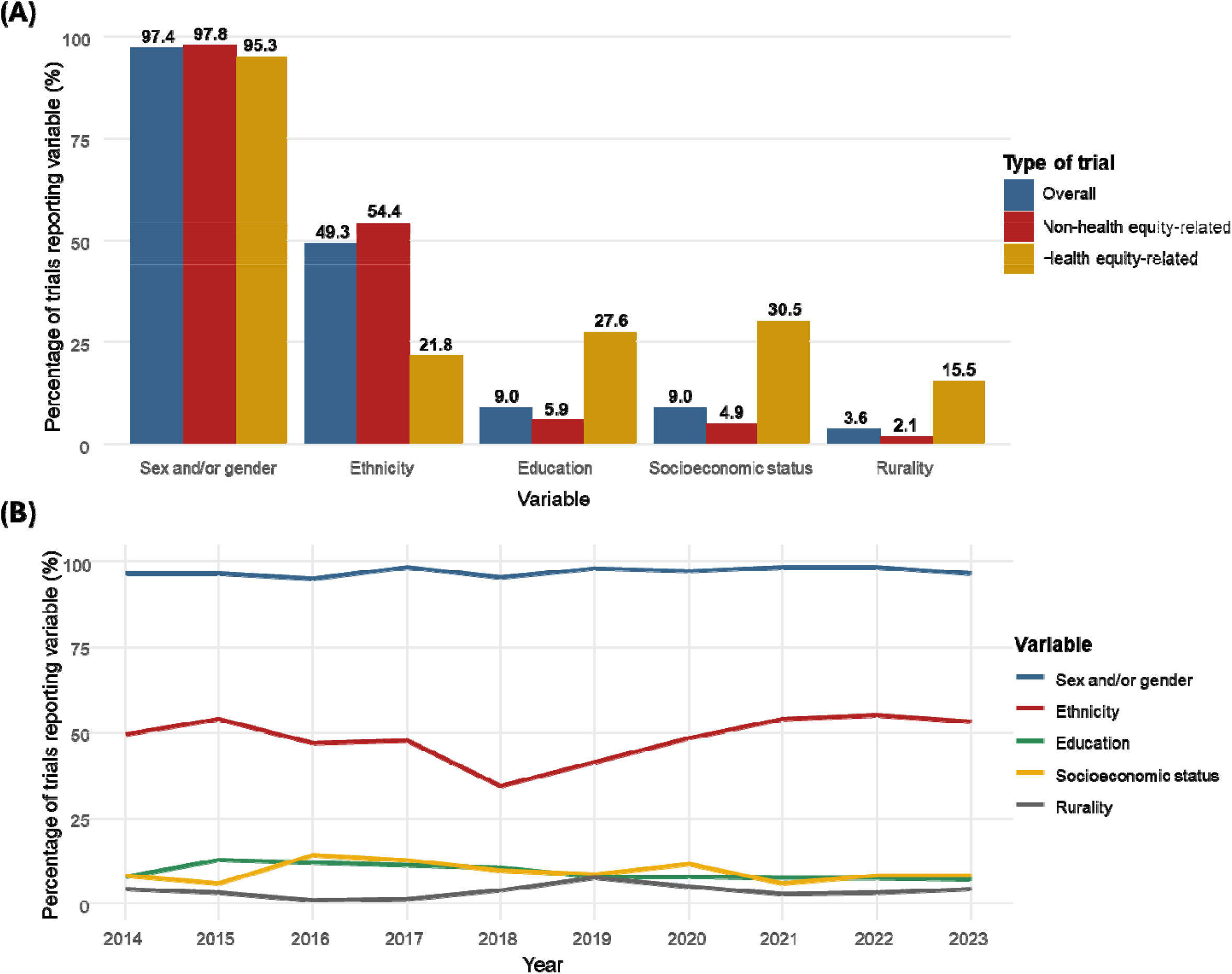
Reporting of each socio-demographic characteristic of interest overall and by trial type (A), and by year (B).

Health equity-related trials were less likely to report ethnicity (21.8% vs 54.4%; *p*<0.0001), but more likely to report education level (27.6% vs 5.9%; *p*<0.0001), socioeconomic status (30.5% vs 4.9%; *p*<0.0001), and rurality (15.5% vs 2.1%; *p*<0.0001) (**Figure 2A**). Reporting of all sociodemographic characteristics did not improve throughout the 10-year study period (**Figure 2B**).

Reporting of each characteristic differed depending on trial subject area (**Figure 3**). Trials studying viral infections, HIV/STIs, and COVID-19 had the highest reporting of ethnicity (69.6%, 62.0%, and 61.6% respectively). Conversely, trials studying tropical medicine, critical care, and antimicrobial stewardship/infection control had the lowest reporting of ethnicity (19.5%, 21.6%, and 21.6% respectively). HIV/STI trials also had the highest reporting of education level (24.6%) and socioeconomic status (17.4%). Trials studying tropical medicine had the highest reporting of rurality/geographical location (11.2%).

**Figure 3:**
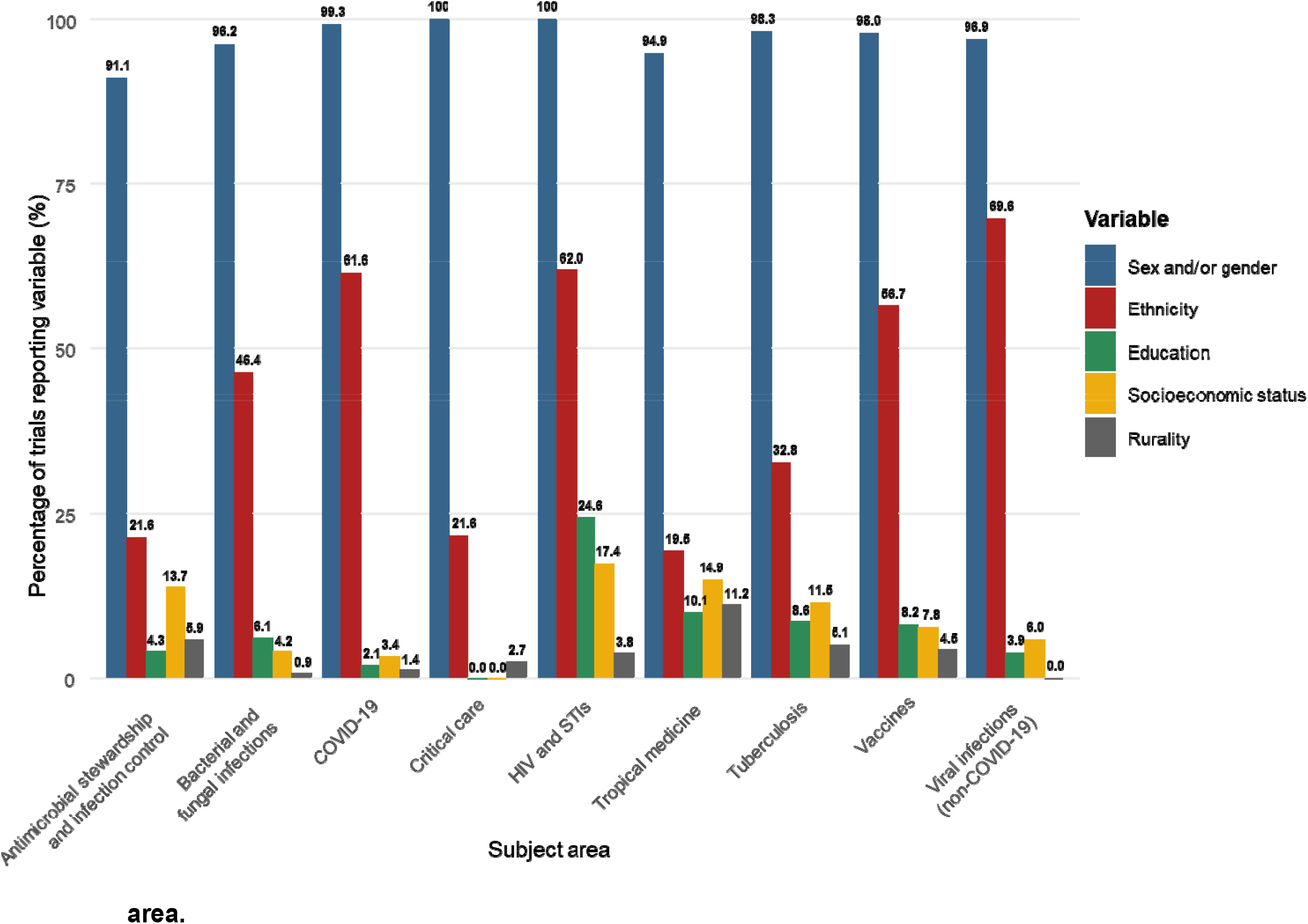
Reporting of each socio-demographic characteristic of interest, by subject area.

Multivariable logistic regression models identified several trial characteristics that were independently associated with reporting of ethnicity, education level, and socio-economic status (**Table 2**). Models for the reporting of sex/gender and rurality/geographical location were not fit because of almost universal reporting in the former and almost universal non-reporting in the latter. Effects of subject area were varied; for example, COVID-19 trials were more likely to report ethnicity but less likely to report education level (compared to the referent group of bacterial/fungal infections). HIV and STI trials were associated with an increased likelihood of reporting of ethnicity, education level, and socio-economic status. Having industry funding was associated with an increased adjusted odds of reporting ethnicity (aOR 2.43, 95% CI 1.60–3.68) but lower adjusted odds of reporting education level (aOR 0.056, 95% CI 0.007–0.43) or socio-economic status (aOR 0.22, 95% CI 0.065–0.77). Health-equity related trials were associated with increased adjusted odds of reporting education level (aOR 3.72, 95% CI 2.14–6.46) and socio-economic status (aOR 5.87, 95% CI 3.46–9.93), but not ethnicity (aOR 0.78, 95% CI 0.49–1.26). Use of a medical writer was associated with increased adjusted odds of reporting ethnicity (aOR 1.79, 95% CI 1.15–2.77) and lower adjusted odds of socio-economic status (aOR 0.20, 95% CI 0.045–0.87). Lastly, trials conducted in lower or middle-income countries were less likely to report ethnicity (aOR 0.15, 95% CI 0.088–0.19) but more likely to report education level (aOR 2.74, 95% CI 1.56– 4.83).

**Table 2:**
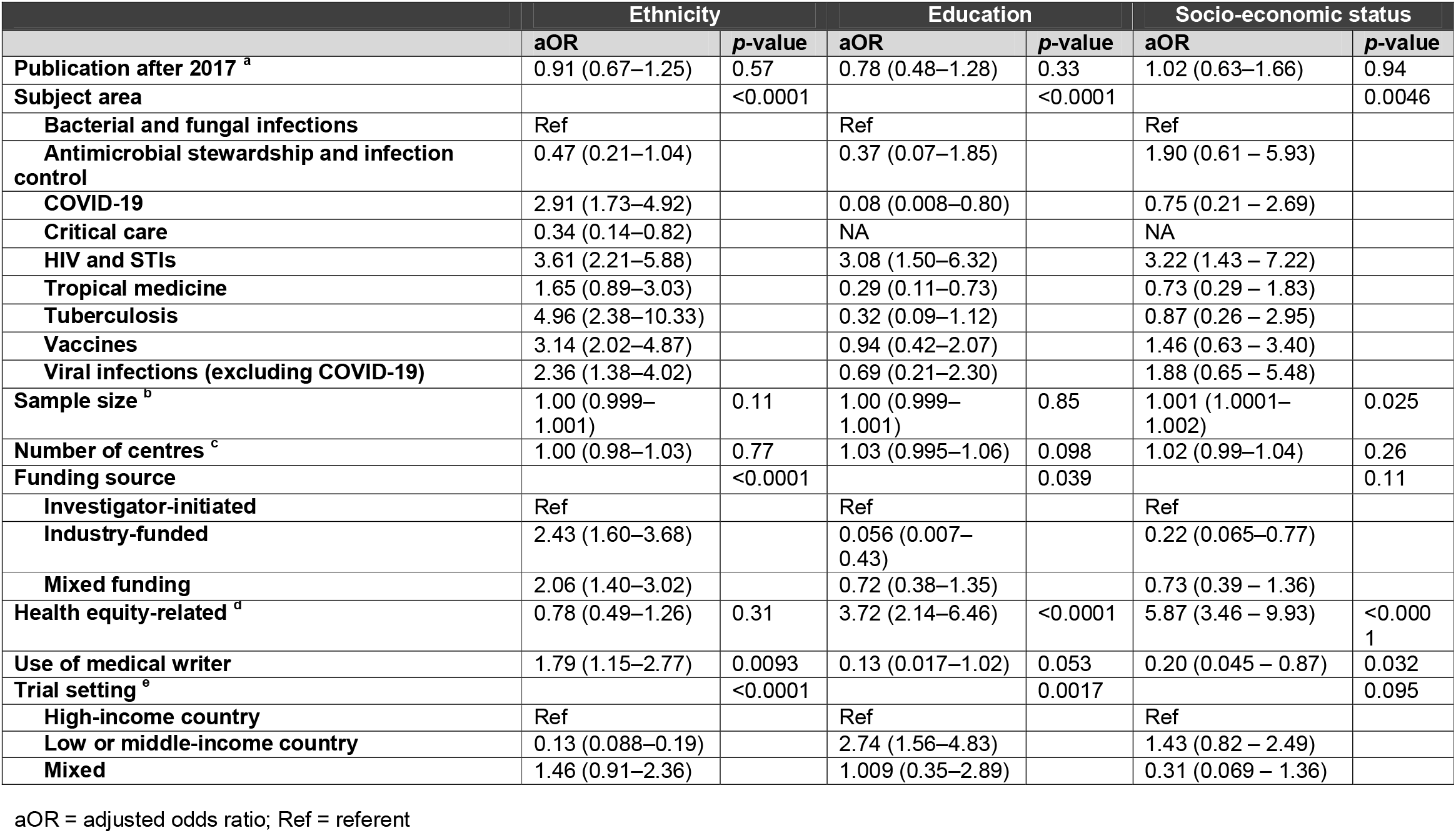
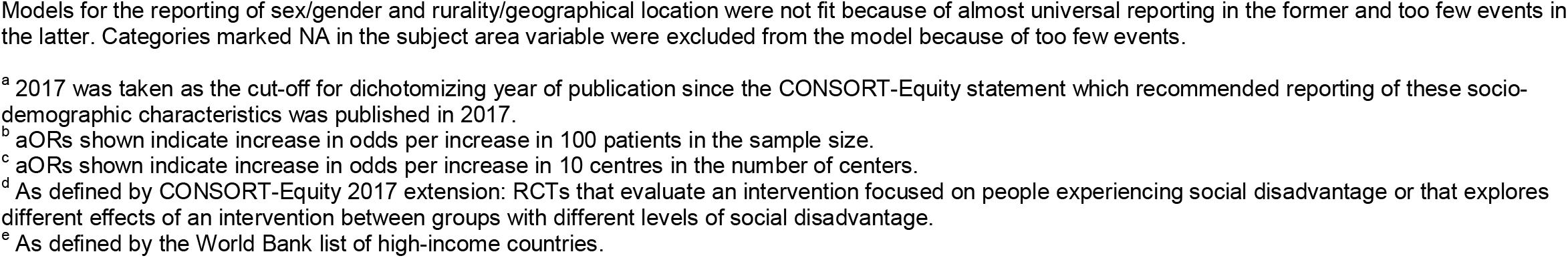
Multivariable logistic regression models identifying trial characteristics associated with reporting of ethnicity, education, and socio-economic status.

|  | Ethnicity |  | Education |  | Socio-economic status |  |
| --- | --- | --- | --- | --- | --- | --- |
|  | aOR | p-value | aOR | p-value | aOR | p-value |
| Publication after 2017 <sup>a</sup> | 0.91 (0.67–1.25) | 0.57 | 0.78 (0.48–1.28) | 0.33 | 1.02 (0.63–1.66) | 0.94 |
| Subject area |  | <0.0001 |  | <0.0001 |  | 0.0046 |
| Bacterial and fungal infections | Ref |  | Ref |  | Ref |  |
| Antimicrobial stewardship and infection control | 0.47 (0.21–1.04) |  | 0.37 (0.07–1.85) |  | 1.90 (0.61 – 5.93) |  |
| COVID-19 | 2.91 (1.73–4.92) |  | 0.08 (0.008–0.80) |  | 0.75 (0.21 – 2.69) |  |
| Critical care | 0.34 (0.14–0.82) |  | NA |  | NA |  |
| HIV and STIs | 3.61 (2.21–5.88) |  | 3.08 (1.50–6.32) |  | 3.22 (1.43 – 7.22) |  |
| Tropical medicine | 1.65 (0.89–3.03) |  | 0.29 (0.11–0.73) |  | 0.73 (0.29 – 1.83) |  |
| Tuberculosis | 4.96 (2.38–10.33) |  | 0.32 (0.09–1.12) |  | 0.87 (0.26 – 2.95) |  |
| Vaccines | 3.14 (2.02–4.87) |  | 0.94 (0.42–2.07) |  | 1.46 (0.63 – 3.40) |  |
| Viral infections (excluding COVID-19) | 2.36 (1.38–4.02) |  | 0.69 (0.21–2.30) |  | 1.88 (0.65 – 5.48) |  |
| Sample size <sup>b</sup> | 1.00 (0.999–1.001) | 0.11 | 1.00 (0.999–1.001) | 0.85 | 1.001 (1.0001–1.002) | 0.025 |
| Number of centres <sup>c</sup> | 1.00 (0.98–1.03) | 0.77 | 1.03 (0.995–1.06) | 0.098 | 1.02 (0.99–1.04) | 0.26 |
| Funding source |  | <0.0001 |  | 0.039 |  | 0.11 |
| Investigator-initiated | Ref |  | Ref |  | Ref |  |
| Industry-funded | 2.43 (1.60–3.68) |  | 0.056 (0.007–0.43) |  | 0.22 (0.065–0.77) |  |
| Mixed funding | 2.06 (1.40–3.02) |  | 0.72 (0.38–1.35) |  | 0.73 (0.39 – 1.36) |  |
| Health equity-related <sup>d</sup> | 0.78 (0.49–1.26) | 0.31 | 3.72 (2.14–6.46) | <0.0001 | 5.87 (3.46 – 9.93) | <0.0001 |
| Use of medical writer | 1.79 (1.15–2.77) | 0.0093 | 0.13 (0.017–1.02) | 0.053 | 0.20 (0.045 – 0.87) | 0.032 |
| Trial setting <sup>e</sup> |  | <0.0001 |  | 0.0017 |  | 0.095 |
| High-income country | Ref |  | Ref |  | Ref |  |
| Low or middle-income country | 0.13 (0.088–0.19) |  | 2.74 (1.56–4.83) |  | 1.43 (0.82 – 2.49) |  |
| Mixed | 1.46 (0.91–2.36) |  | 1.009 (0.35–2.89) |  | 0.31 (0.069 – 1.36) |  |

aOR = adjusted odds ratio; Ref = referent
Models for the reporting of sex/gender and rurality/geographical location were not fit because of almost universal reporting in the former and too few events in the latter. Categories marked NA in the subject area variable were excluded from the model because of too few events.
<sup>a</sup> 2017 was taken as the cut-off for dichotomizing year of publication since the CONSORT-Equity statement which recommended reporting of these socio-demographic characteristics was published in 2017.
<sup>b</sup> aORs shown indicate increase in odds per increase in 100 patients in the sample size.
<sup>c</sup> aORs shown indicate increase in odds per increase in 10 centres in the number of centers.
<sup>d</sup> As defined by CONSORT-Equity 2017 extension: RCTs that evaluate an intervention focused on people experiencing social disadvantage or that explores different effects of an intervention between groups with different levels of social disadvantage.
<sup>e</sup> As defined by the World Bank list of high-income countries.

## DISCUSSION

In this study, we systematically reviewed 1343 infectious disease RCT publications across 10 medical journals over a 10-year period. We found that reporting of socio-demographic variables was low, inconsistent across different trial subject areas, and has not improved over time. Our findings are consistent with similar studies in general medical journals and across multiple medical specialties, which have reported similarly low and inconsistent reporting of these socio-demographic variables, particularly race/ethnicity and socioeconomic status [14-16, 20-24]. In addition, our findings build upon existing data by focusing on a broader range of socio-demographic variables, examining a larger number of papers, and identifying specific trial characteristics associated with increased reporting. To our knowledge, this is also the first report specifically examining the field of infectious diseases, a field where social determinants of health are especially important, given the unique intersection of infectious diseases with inequity, marginalization, and other social issues (e.g., immigration, housing instability, substance use, sexual health, etc.) [25, 26]. Collectively, this body of literature should prompt the clinical research community to develop consensus statements and/or guidelines outlining the minimally required set of socio-demographic variables that should be reported in RCT publications, and describe the specific trial types and/or settings that require more detailed reporting. Several high impact medical journals have published guidance for the reporting of socio-demographic variables in research reports [27, 28], but uptake of and adherence to these recommendations remains inconsistent.

The importance of adequate representation of patients of minority backgrounds or of marginalized status in RCTs has been increasingly recognized [29]. From the researcher’s and scientific community’s perspective, ensuring that trial populations are reflective of the real-world patient populations (including their distribution of socio-demographic characteristics) improves generalizability of trial results, clinicians’ confidence in trial findings, and subsequent translation to clinical practice [30, 31]. From a patient’s perspective, there may be mistrust in trial fundings or medical research from specific population groups, unless they feel that they were well-represented in studies [32]. From an equity and medical ethics perspective, clinical trials are a scarce resource that should be fairly distributed across all segments of the patient population [33, 34]. Limiting research to specific patient subgroups or strata of the patient population may exacerbate existing inequities instead of correcting them. Despite this, data consistently shows that patients of minority ethnic background or of lower socio-economic status remain under-represented in RCTs [23, 35, 36]. Calls are being made to ensure that researchers take active measures to improve representation of these historically under-represented groups in clinical trials [28, 37]. Alongside any interventions, adequate and transparent reporting of socio-demographic characteristics in trial populations is essential to track and assess the impact of these interventions over time.

Beyond tracking absolute reporting (i.e., *if* these variables are being reported), it is equally important to ensure quality and consistency of reporting (i.e., *how* these variables are being reported). In our study, 17.9% of the studies that reported ethnicity only reported binary categories (e.g. white vs non-white), which is likely to be inadequate in capturing the broad diversity of most patient populations. The *American Medical Association Manual of Style* has published guidance on how ethnicity should be reported in research reports [27]. Multiple specific categories should be reported, rather than condensing multiple ethnicity groups into one. Categories may differ depending on the country where a study is conducted, however they should be consistent within studies from the same country to permit comparisons across time and across different studies. It is also important to report how ethnicity data were collected (e.g., self-reported and self-identified by participants, or extracted from an electronic record/database); self-reporting and self-identification of ethnicity should be prioritized where possible. We also describe the many different ways how education level, socioeconomic status, and rurality variables were reported. Similar guidance should be developed by professional bodies to ensure consistency in the reporting of these variables.

There are several limitations to our study. Firstly, due to resource limitations, it was unfeasible to review all published infectious disease related RCTs during the 10-year study period without additional restrictions. We thus chose to focus on 10 journals that frequently publish RCTs. Our findings may thus not be reflective of the entire body of infectious disease literature. Nonetheless, as we prioritized high impact journals with wide readership (several of which have editorial policies emphasizing the importance of reporting of these socio-demographic variables), it is likely that these journals have a higher-than-average frequency and quality of reporting. Expansion of our study without journal limitations is unlikely to find an improvement in the frequency of reporting of these variables. Secondly, we focused on assessing if and how RCTs reported different socio-demographic variables, but did not assess whether the enrolled populations in each RCT were representative of their broader patient populations, which is beyond the scope of this study.

## CONCLUSION

Reporting of socio-demographic characteristics in infectious diseases RCTs is inconsistent and has not improved over time. Further efforts should be made by the research community and professional bodies to improve reporting standards and ensure consistency of reporting of these variables, with the broader aim of improving diverse representation in RCTs and equitable access to research opportunities for patients.

## Data Availability

All data produced in the present study are available upon reasonable request to the corresponding author.

## Ethical approvals

No ethics approvals or informed consent was required for the conduct of this systematic review.

