## Supplementary Appendix for "Reporting of socio-demographic characteristics of trial participants in infectious diseases clinical trials – a systematic review"

**Contents:**


1. Supplementary methods
2. Search strategy
3. Supplementary Table 1: Details of reporting of each socio-demographic variable

### 1. Supplementary Methods

*Definitions*

Ethnicity: ethnicity refers to groups of people whose members considers themselves distinct within a society, and could be due to shared origin/background, culture, traditions, or language. “Race” is often used interchangeably but is more controversial as its use may imply natural or biological differences which may not be grounded in scientific evidence.[10] For this systematic review, either term (or other related terms, e.g., “ethnic background”, “racialized group”) was considered as presence of reporting.

Sex and/or gender: sex refers to biological attributes of an individual, often categorized in a binary as female or male. Gender refers to the socially constructed roles, behaviours, expressions, and identities, and is not confined to a binary.

Education level: this can be reported in several ways, e.g., highest level of education attained (categorical), number of years of formal schooling (integer), completion of secondary school (binary). Any of these was considered as presence of reporting.

Socio-economic status: this can be reported in terms of income level, employment status, or other measurement indices derived from census data (e.g., the Ontario marginalization index).

Rurality or geographical location: geographical location is an important determinant of health and can reported in many ways depending on setting and context. For example, it can reported in a binary manner (remote vs non-remote), categorical manner (urban, inner city, rural), or quantitative manner (rurality index, distance from nearest health centre or hospital). Any of these was considered as presence of reporting.

### 2. Search Strategy

| | |
|---|---|
| 1 | exp Infections/ |
| 2 | ((randomized controlled trial or controlled clinical trial).pt. or randomized.ab. or placebo.ab. or clinical trials as topic.sh. or randomly.ab. or trial.ti. or clinical trial.mp. or clinical trial.pt. or random*.mp.) not (animals not (humans and animals)).sh. |
| 3 | 1 and 2 |
| 4 | limit 3 to (english language and full text and yr="2013 - 2023") |
| 5 | ("new england journal of medicine" or jama or "british medical journal" or lancet or "annals of internal medicine" or "clinical infectious diseases" or "lancet infectious diseases" or "clinical microbiology & infection" or "journal of infection" or "journal of infectious diseases").jn. |
| 6 | 4 and 5 |

#### 3. Supplementary Table 1: Details of reporting of each socio-demographic variable.

| **Variable** | **Number (%)** |
|---|---|
| Ethnicity | 654 of 1326 (49.3%) |
| Binary (only two categories stated, e.g. white/non-white | 117 of 654 (17.9%) |
| Categorical (multiple categories stated) | 537 of 654 (82.1%) |
| Education level | 113 of 1252 (9.0%) |
| Categorical (highest education level reached) | 65 of 113 (57.5%) |
| Binary (completion of specific milestone, e.g. secondary school) | 31 of 113 (27.4%) |
| Number of years of schooling | 12 of 113 (10.6%) |
| Others (e.g., health literacy level, ability to read and write) | 5 of 113 (4.4%) |
| Socioeconomic status | 120 of 1340 (9.0%) |
| Employment status | 41 (34.2%) |
| Categorical (different income levels) | 31 (25.8%) |
| Housing status / type | 16 (13.3%) |
| Binary (above/below specified income cut-off) | 10 (8.3%) |
| Summary index (e.g. marginalization index, SEIFA score) | 10 (8.3%) |
| Insurance status | 6 (5.0%) |
| Absolute income | 6 (5.0%) |
| Occupation | 5 (4.2%) |
| Rurality or geographical location | 45 of 1269 (3.9%) |
| Binary (remote vs non-remote) | 20 (44.4%) |
| Categorical (e.g. urban, inner city, rural) | 10 (22.2% |
| Distance from nearest health centre of hospital | 8 (17.8%) |
| Others (e.g. visual/map depiction, rurality index) | 7 (15.6%) |

SEIFA = socio-economic index for area
